# SMORASO-DT : A hybrid machine learning classification model to classify individuals based on working memory load in mental arithmetic task

**DOI:** 10.1101/2020.10.02.20205922

**Authors:** K R Shivabalan, Brototo Deb, Shivam Goel, R Arivan

## Abstract

Nonlinear dynamics and chaos theory are being widely used nowadays in neuroscience to characterize complex systems within which the change of the output is not proportional to the change applied at the input. Such nonlinear systems compared to linear systems, often appear chaotic, unpredictable, or counterintuitive, however, yet their behaviour is not mapped out as random. Thus, hidden potential of the dynamical properties of the physiological phenomenon can be detected by these approaches especially to elucidate the complex human brain activity gathered from the electroencephalographic (EEG) signals. As it is known, brain is a chaotic dynamical system and its generated EEG signals are generally chaotic because, with respect to time, the amplitude changes continuously. A reliable and non-invasive measurement of memory load, to measure continuously while performing a cognitive task, is highly desirable to assess cognitive functions, crucial for prevention of decision-making errors. Such measurements help to keep up the efficiency and productivity in task completion, work performance, and to avoid cognitive overload, especially at high mental or physical workload places like traffic control, military operations, and rescue commands. In this work, we have measured the linear and nonlinear dynamics of the EEG signals in subjects undergoing mental arithmetic task. Further, we have also differentiated the subjects who can perform a mental task good or bad, and developed a hybrid machine learning model, the SMORASO-DT (SMOte + Random forest + lASso-Decision Tree), to differentiate good and bad performers during n-back task state with an accuracy rate of 78%.

## Introduction

Complex task performance requires the integration of knowledge related to the task, working memory, attention and decision making. Cognitive load refers to the amount of mental demand imposed by a particular task on these parameters^1^. A reliable, objective and cost-effective method of assessing cognitive load is crucial in understanding the mental capacity of people. Experts have more knowledge or experience with regards to a specific task which reduces the cognitive load associated with the task in contrast to novices who have an elevated cognitive load^2^. Measuring and assessing the cognitive load associated with different tasks is crucial for many applications, including monitoring the mental well-being of people who require heavy cognitive load in their occupation. This load increases the chances of error in the task at hand^3–5^ or stereotyping^6^. Subjective measures of cognitive load^4^ have been useful for cognitive load measurement in embodied scenarios, in cases where an appropriate survey is chosen. However, the information gathered via these routes is limited since the use of such cognitive load questionnaires have their own theoretical and practical issues. Moreover, use of different phrases in cognitive load questionnaires may lead to results that are not comparable^7^. In addition to behavioural aforementioned measures, cognitive workload has been assessed by physiological measures, such as measurement of pupil diameter^8^. The application of objectively determining the cognitive load has its growing application in ergonomics among the pilots or drivers.^9^

In this context, gathering electroencephalography (EEG) data is an easy, cost effective investigation to assess the brain’s activity. The focus of this work is, thus, to quantify cognitive workload using measures based on EEG data. Recently, EEG-based measures for cognitive load have led to a resurgence of interest in several studies. In fact, this relationship among different spectral features for predicting cognitive load from EEG has been described in many studies^10–12^.

In order to leverage the rich is information in linear and non-linear dynamic features of complex signals such as the EEG, a wide range of methods have been applied for measurement and classification of the memory load using EEG signal. Another reason behind our preference to utilize nonlinear measures to analyse EEG data is that nonlinear (non-stationary and irregular) signals nullify comprehensive understanding by a classic reductionist approach. Even the simplest nonlinear signals (originating from complex systems) nullify the criteria of proportionality and superposition characteristics for linear systems^13^. The application of such non-linear methods in classifying mental tasks is quite recent, and measures like correlation dimension (CD)^14–16^, fractal dimension^17–19^, Hurst exponent (HE)^9,15,20^, sample entropy^18^, approximate entropy (ApEn)^15,21,22^ and largest Lyapunov exponent (LLE)^14,15^ have been used to measure the complexity/irregularity of the underlying brain dynamics during its performance of some cognitive tasks compared to the rest condition. In other words, in these studies the brain activity states: such as the rest and stimulated states have been differentiated. But to date, these measures have not been investigated in the analysis of the varying working memory load and the question whether these approaches could provide some information on discriminating subjects who can perform the arithmetic task well and subjects who cannot. In future, this study can be potentially extrapolated to subjects with dementia or minimal cognitive impairment and will be able to detect such degenerative processes at earlier stages and can help to understand the progression of these diseases as far as electrophysiology is concerned. This study also ensures productivity in task completion and applications in ergonomics.

## Results

### 1. EEG Feature extraction

First we extracted the linear and non linear features of EEG signal of all leads in both good and bad groups of the dataset using the EEGFrame software. Then the groups are classified using random forest algorithm implemented in R.

### 2. Feature selection: Random forest

#### A. Distribution of minimal depth

**Fig. 1.**
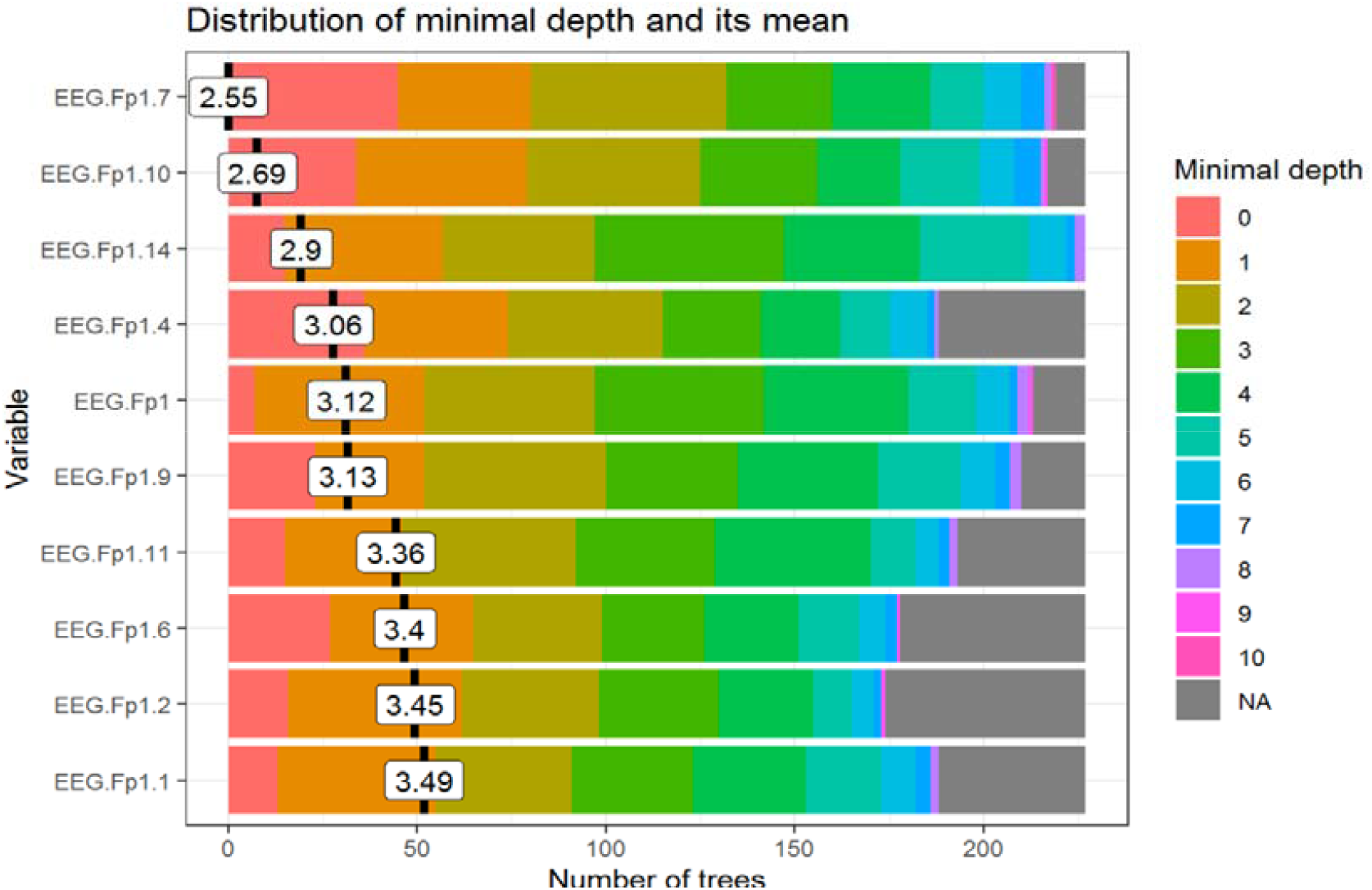
Plot showing the distribution of minimal depth among the trees of the forest. X axis scale goes from zero to the maximum number of trees in which any variable was used for splitting. Y axis showing the mean of the distribution with a value label on it. Minimal depth for a variable in a tree equals to the depth of the node which splits on that variable and is closest to the root of the tree. Each linear or non linear measure has been given a code name which has been added in the supplementary file. For example, EEG Fp1.7 is approximate entropy of the Fp1 signal.

#### B. Multi-way importance plot

The multi-way importance plot shows the relation between three measures of importance and labels 10 variables which scored best when it comes to these three measures (i.e. for which the sum of the ranks for those measures is the lowest).

**Fig. 2.**
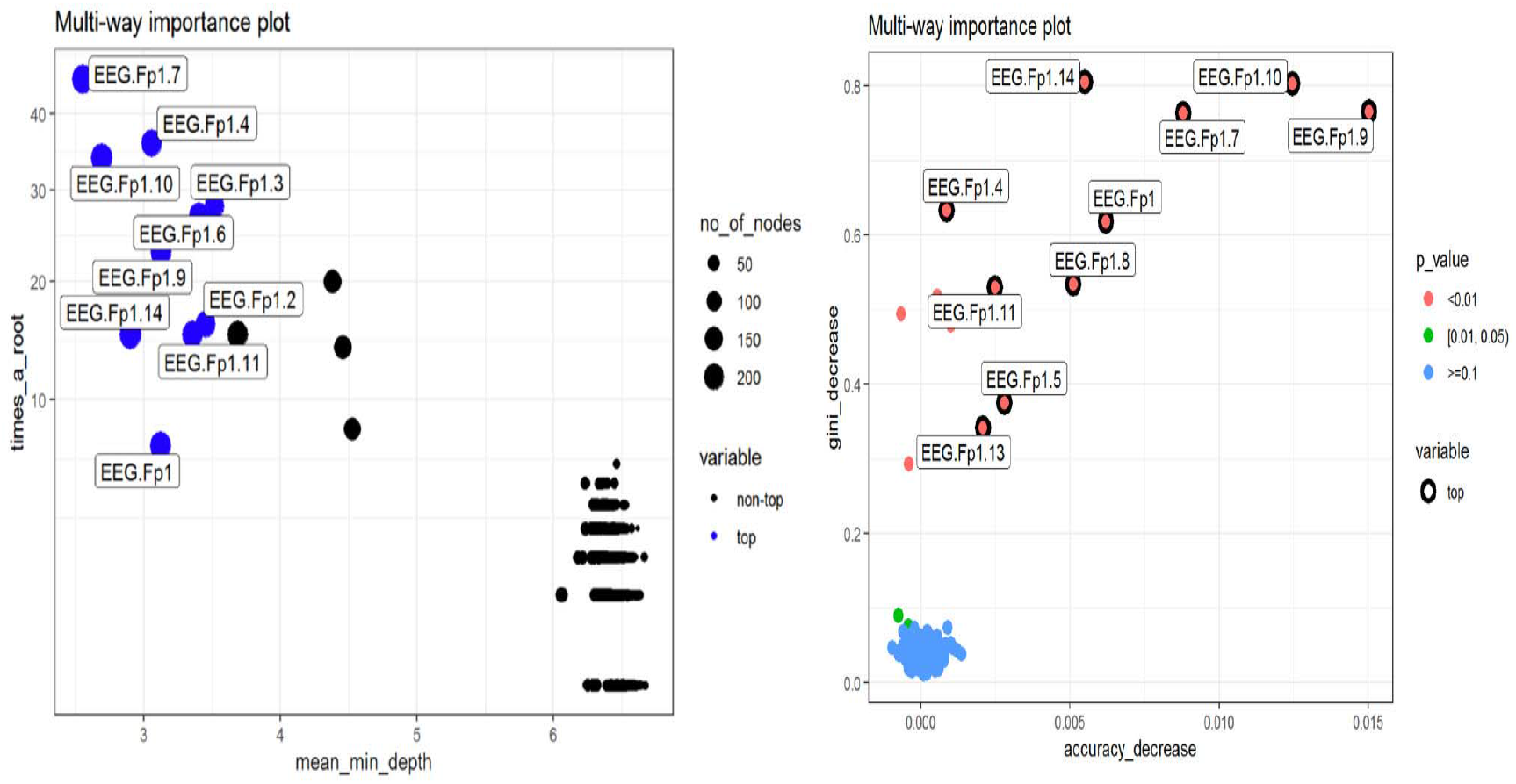
The first multi-way importance plot focuses on three importance measures that derive from the structure of trees in the forest: mean depth of first split on the variable, number of trees in which the root is split on the variable, the total number of nodes in the forest that split on that variable. The second multi-way importance plot shows importance measures that derive from the role a variable plays in prediction: accuracy_decrease and gini_decrease with the additional information on the - value based on a binomial distribution of the number of nodes split on the variable assuming that variables are randomly drawn to form splits (i.e. if a variable is significant it means that the variable is used for splitting more often than would be the case if the selection was random).

**Fig. 3.**
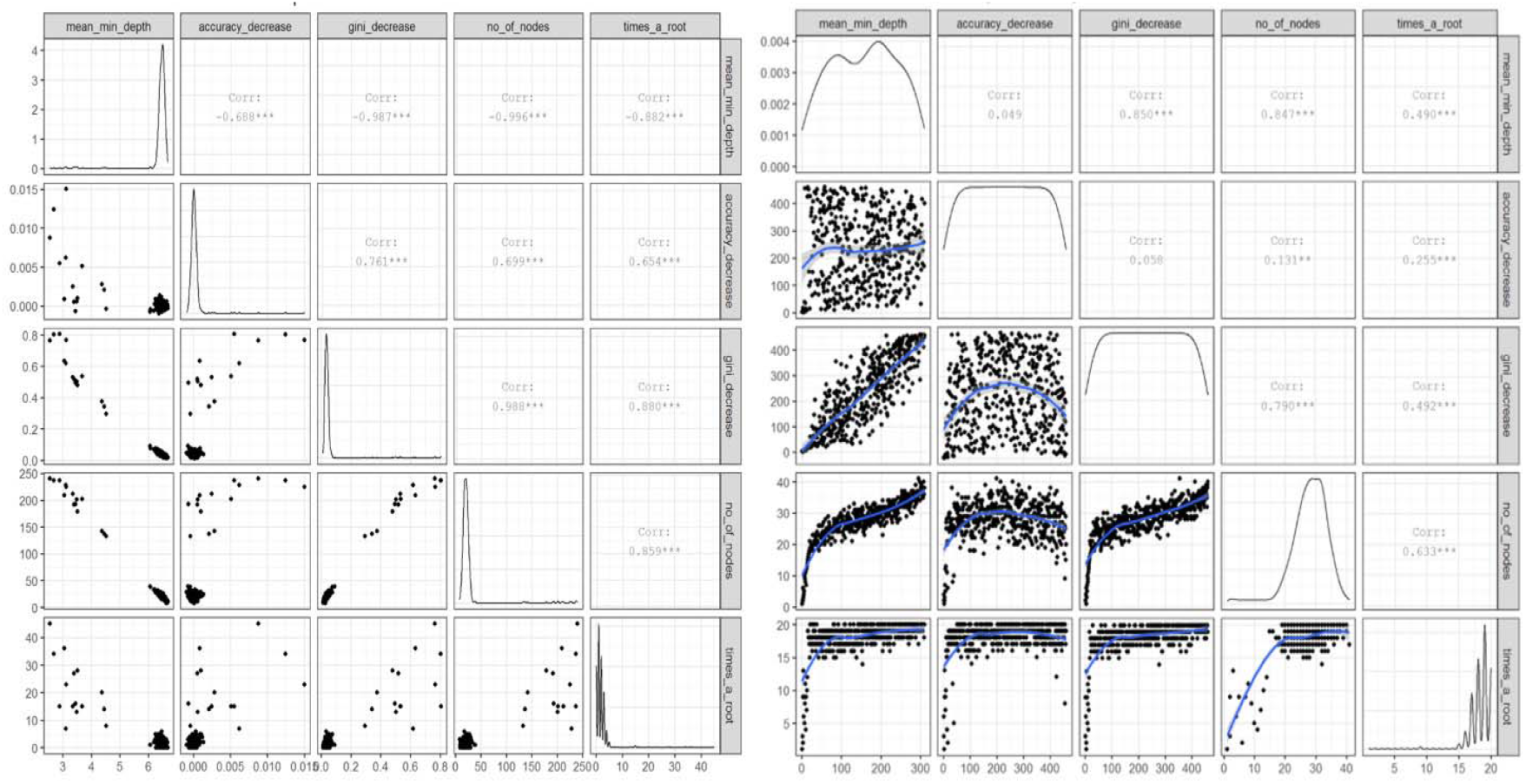
The first plot shows bilateral relations between the following importance measures: mean_min_depth, accuracy_decrease, gini_decrease, no_of_nodes, times_a_root, if some variables are strongly related to each other it may be worth to consider focusing only on one of them. The second plot shows bilateral relations between the rankings of variables according to chosen importance measures. This approach might be useful as rankings are more evenly spread than corresponding importance measures. This may also more clearly show where the different measures of importance disagree or agree.

#### C. Variable interactions

##### a. Conditional minimal depth

The plot below reports 30 top interactions according to mean of conditional minimal depth – a generalization of minimal depth that measures the depth of the second variable in a tree of which the first variable is a root (a subtree of a tree from the forest). In order to be comparable to normal minimal depth 1 is subtracted so that 0 is the minimum. For example, value of 0 for interaction x:y in a tree means that if we take the highest subtree with the root splitting on x then y is used for splitting immediately after x (minimal depth of x in this subtree is 1). The values presented are means over all trees in the forest.

**Fig. 4.**
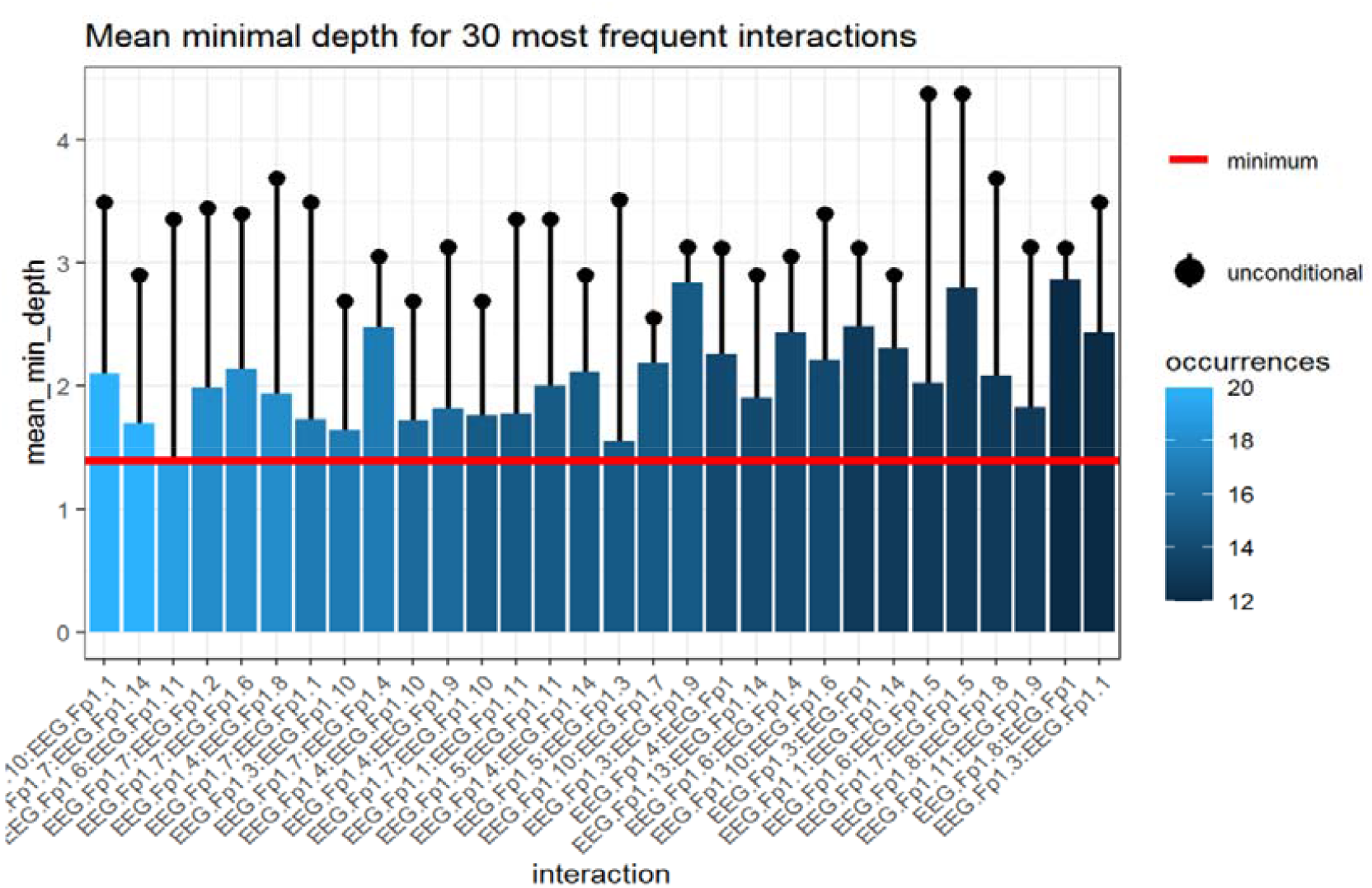
The plot shows only 30 interactions that appeared most frequently, the horizontal line shows the minimal value of the depicted statistic among interactions for which it was calculated, the interactions considered are ones with the following variables as first (root variables): EEG.Fp1.10, EEG.Fp1.7, EEG.Fp1.14, EEG.Fp1.9, EEG.Fp1, EEG.Fp1.11, EEG.Fp1.4, EEG.Fp1.8, EEG.Fp1.5, EEG.Fp1.3, EEG.Fp1.13, EEG.Fp1.1, EEG.Fp1.6, EEG.Fp1.17, EEG.C3.6 and all possible values of the second variable.

##### b. Prediction on a grid

The plots below show predictions of the random forest depending on values of components of an interaction (the values of remaining predictors are sampled from their empirical distribution) for up to 3 most frequent interactions that consist of two numerical variables.

**Fig. 5.**
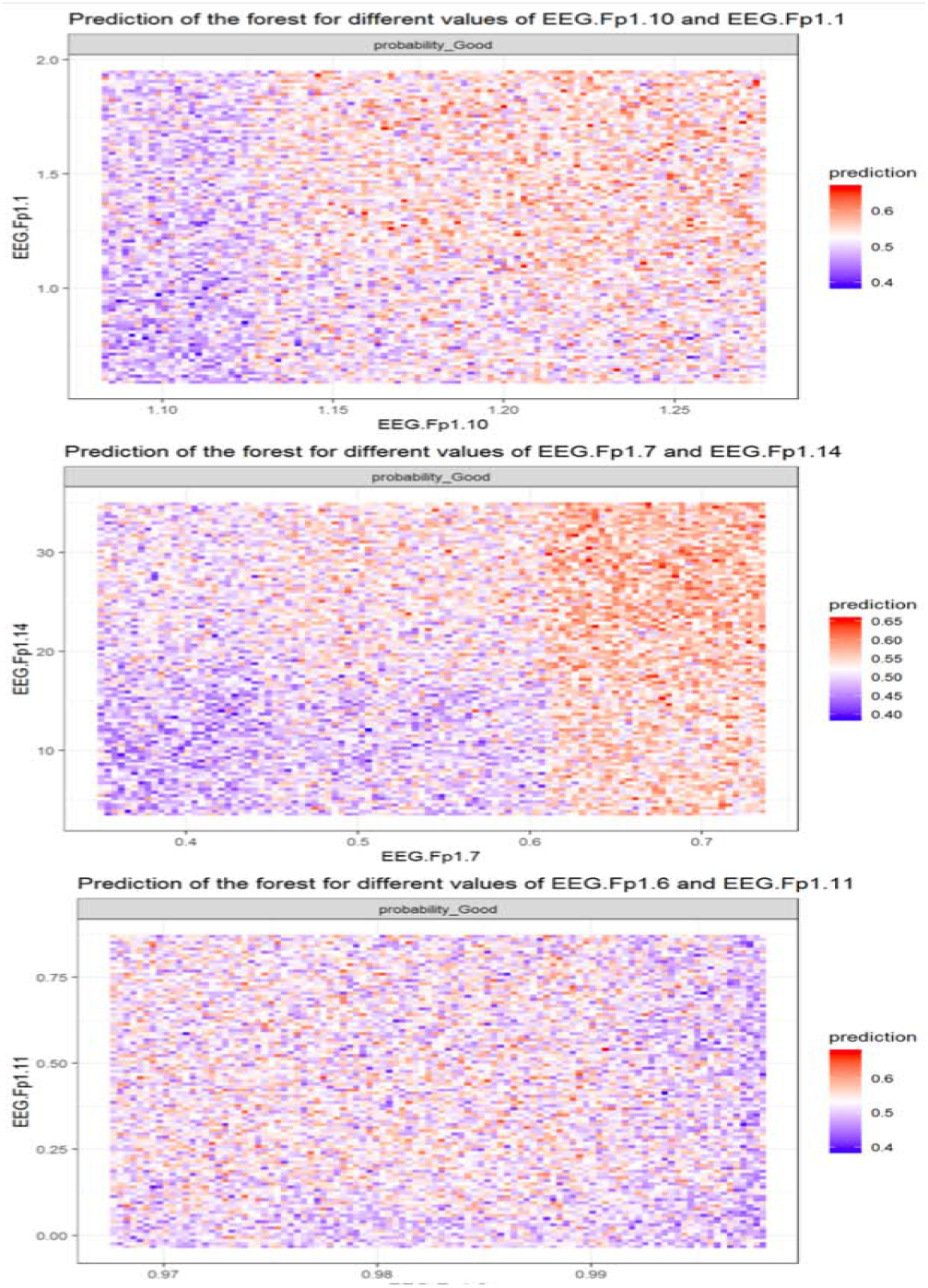
Interaction heatmap showing the prediction of the forest for the different values of the variables.

### 3. Adaptive Lasso with AICc Validation

The top 10 features extracted from random forest are then further made to undergo feature selection using ALASSO.

**Table 1.**
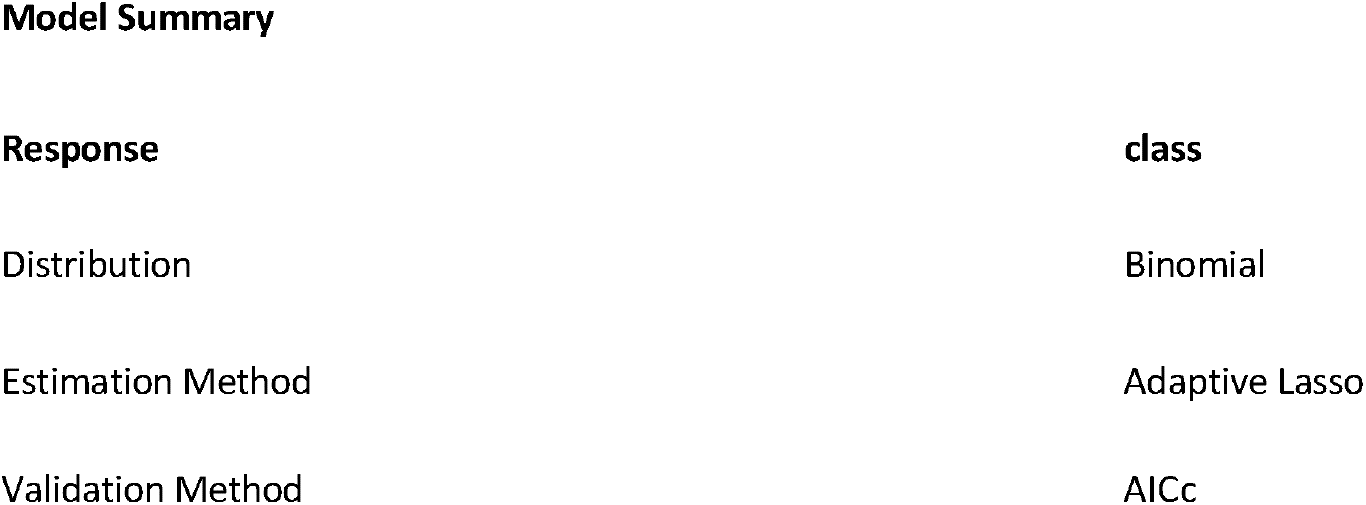
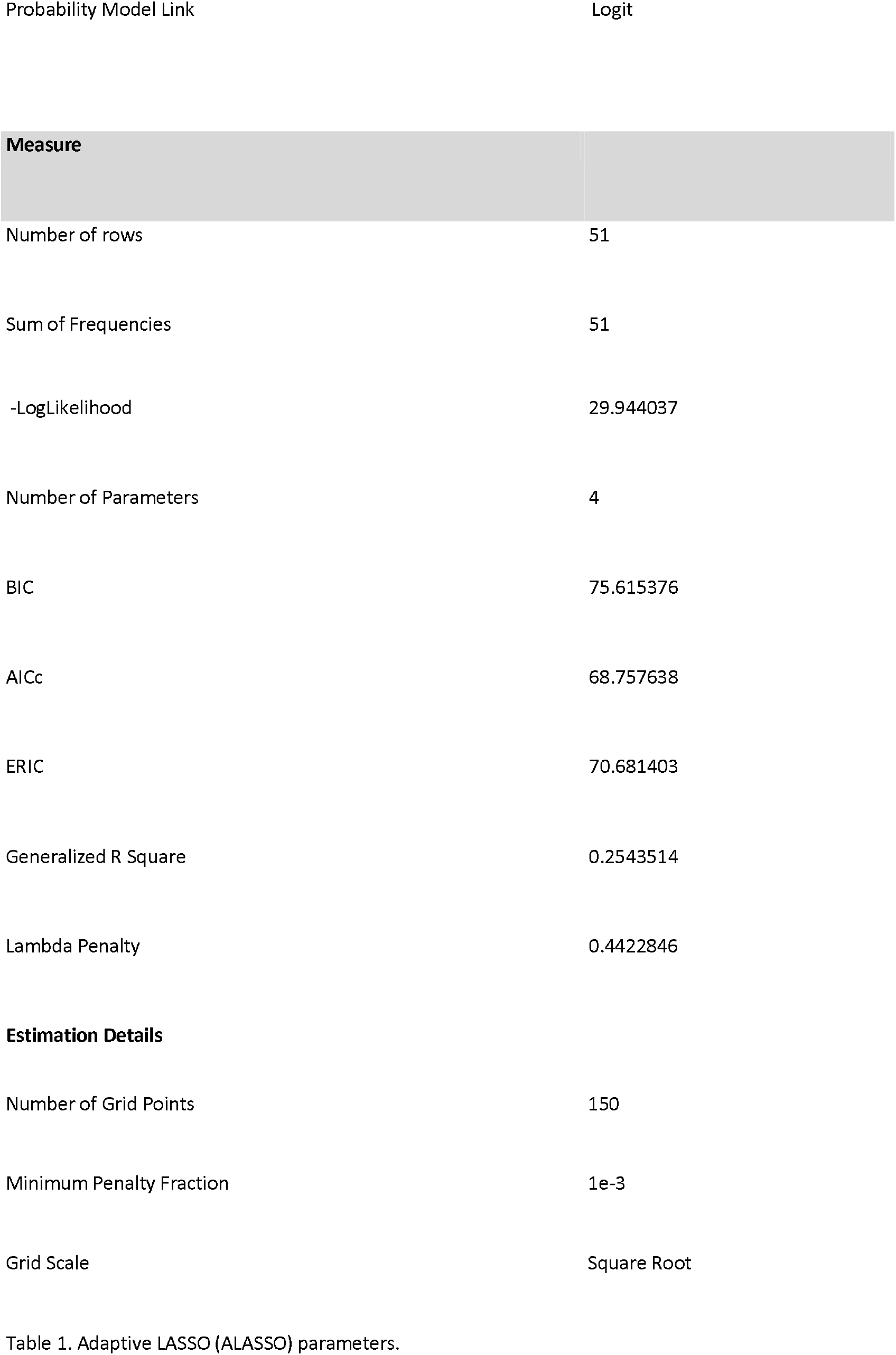
Adaptive LASSO (ALASSO) parameters.

**Figure. 6.**
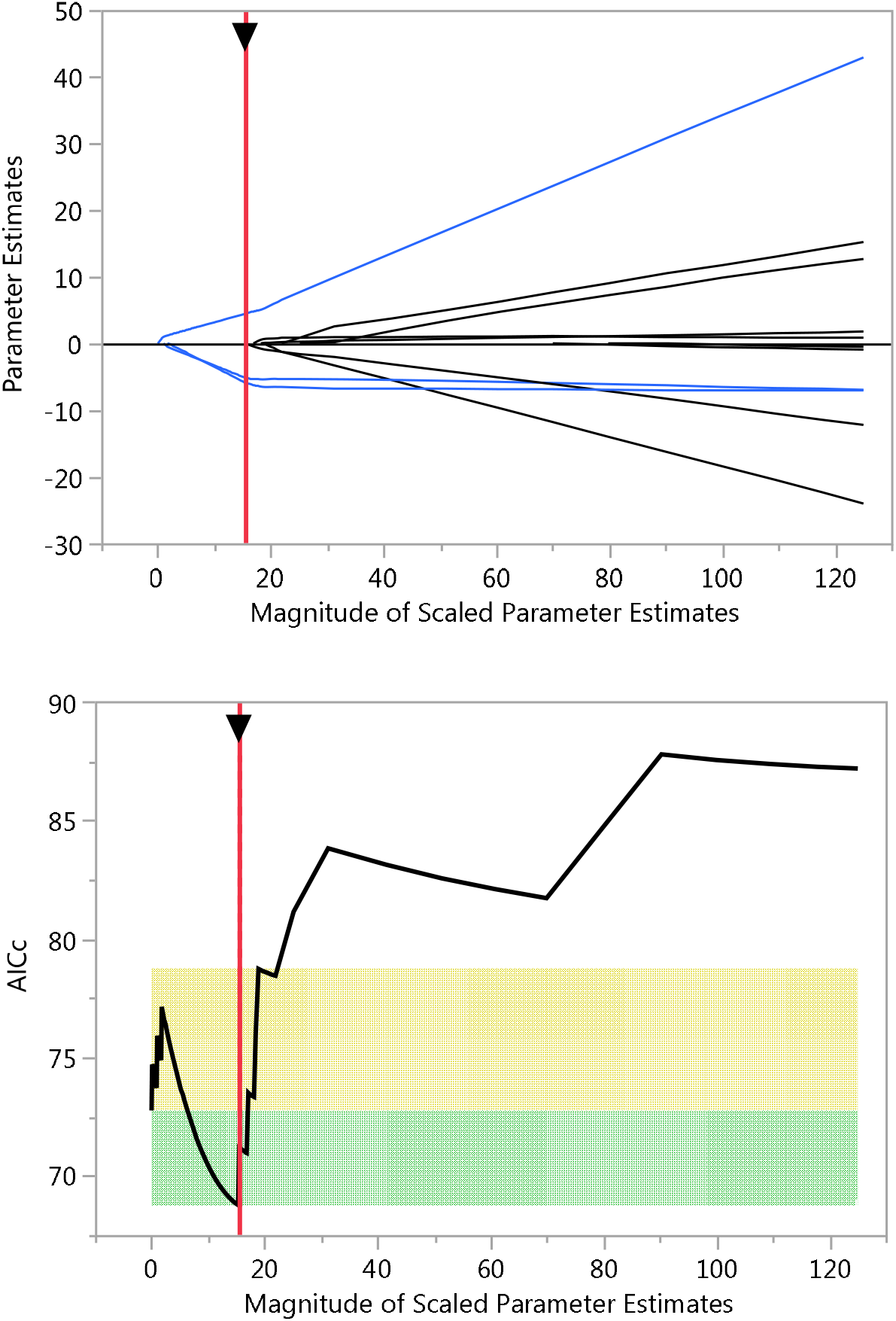
Magnitude of the scaled parameter estimate against the parameter estimates and A1Cc values.

The intercept, EEG.Fp1.5, 10, 14 showed statistically significant results as shown in table 2.

**Table 2.** Parameter Estimates for Original Predictors

| Term | Estimate | Std Error | Wald ChiSquare | Prob > ChiSquare | Lower 95% | Upper 95% |
| --- | --- | --- | --- | --- | --- | --- |
| Intercept | 20.32205 | 9.8453147 | 4.2606495 | 0.0390* | 1.0255874 | 39.618512 |
| EEG.Fp1 | 0 | 0 | 0 | 1.0000 | 0 | 0 |
| EEG.Fp1.1 | 0 | 0 | 0 | 1.0000 | 0 | 0 |
| EEG.Fp1.2 | 0 | 0 | 0 | 1.0000 | 0 | 0 |
| EEG.Fp1.3 | 0 | 0 | 0 | 1.0000 | 0 | 0 |
| EEG.Fp1.4 | 0 | 0 | 0 | 1.0000 | 0 | 0 |
| EEG.Fp1.5 | 0.1782498 | 0.0895754 | 3.9598695 | 0.0466* | 0.0026853 | 0.3538143 |
| EEG.Fp1.7 | 0 | 0 | 0 | 1.0000 | 0 | 0 |
| EEG.Fp1.8 | 0 | 0 | 0 | 1.0000 | 0 | 0 |
| EEG.Fp1.9 | 0 | 0 | 0 | 1.0000 | 0 | 0 |
| EEG.Fp1.10 | -17.95643 | 8.0508523 | 4.9745782 | 0.0257* | -33.73581 | -2.177049 |
| EEG.Fp1.11 | 0 | 0 | 0 | 1.0000 | 0 | 0 |
| EEG.Fp1.12 | 0 | 0 | 0 | 1.0000 | 0 | 0 |
| EEG.Fp1.13 | 0 | 0 | 0 | 1.0000 | 0 | 0 |
| EEG.Fp1.14 | -0.123209 | 0.0560068 | 4.8395472 | 0.0278* | -0.232981 | -0.013438 |
| EEG.Fp1.15 | 0 | 0 | 0 | 1.0000 | 0 | 0 |

#### ROC Curve for class = bad

The AUROC for training of the bad class is around 74.3%.

#### Training

**Figure 7.**
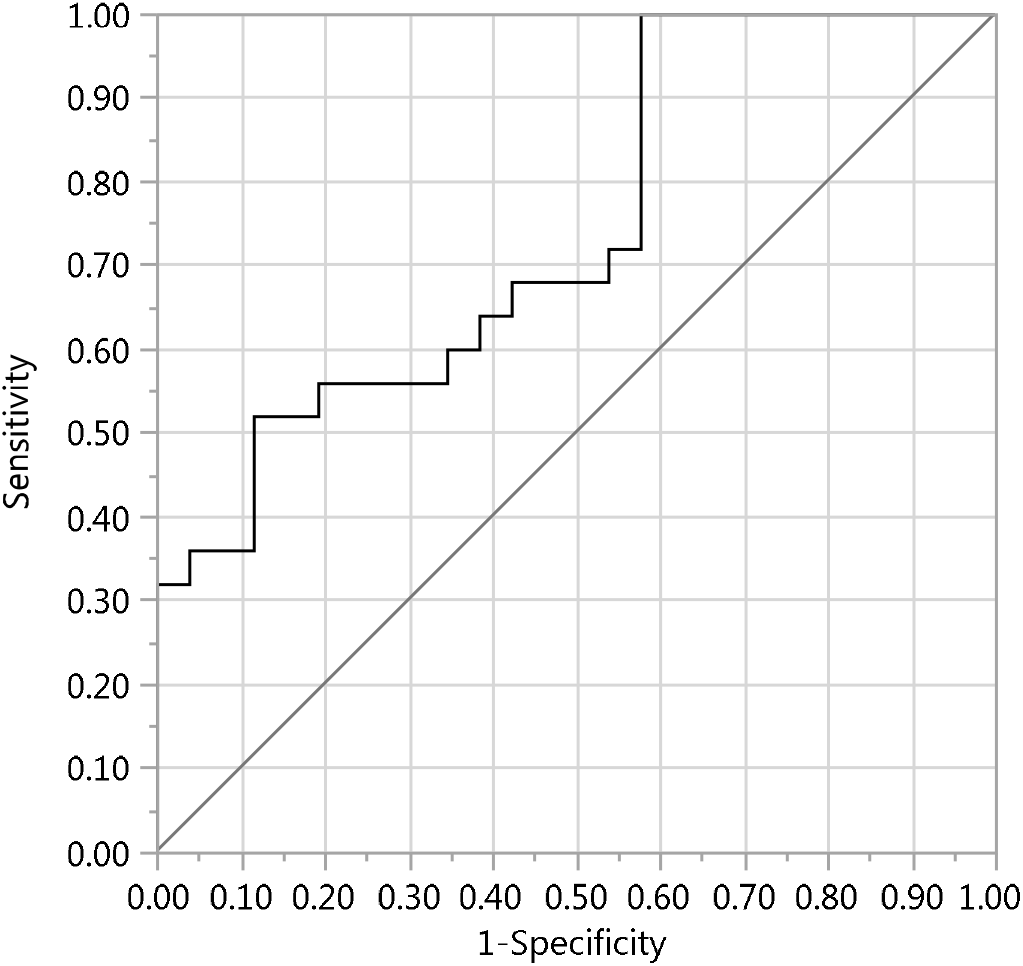
AUROC of 74.3% obtained by training.

#### Odds Ratios

Tests and confidence intervals are Wald based.

Per unit change in regressor is as follows.

**Table 3.** Unit Odds Ratios

| Term | Odds Ratio | Lower 95% | Upper 95% |
| --- | --- | --- | --- |
| EEG.Fp1 | 1 | 1 | 1 |
| EEG.Fp1.1 | 1 | 1 | 1 |
| EEG.Fp1.2 | 1 | 1 | 1 |
| EEG.Fp1.3 | 1 | 1 | 1 |
| EEG.Fp1.4 | 1 | 1 | 1 |
| EEG.Fp1.5 | 1.1951238 | 1.0026889 | 1.4244906 |
| EEG.Fp1.7 | 1 | 1 | 1 |
| EEG.Fp1.8 | 1 | 1 | 1 |
| EEG.Fp1.9 | 1 | 1 | 1 |
| EEG.Fp1.10 | 1.5908e-8 | 2.232e-15 | 0.1133756 |
| EEG.Fp1.11 | 1 | 1 | 1 |
| EEG.Fp1.12 | 1 | 1 | 1 |
| EEG.Fp1.13 | 1 | 1 | 1 |
| EEG.Fp1.14 | 0.8840786 | 0.7921689 | 0.986652 |
| EEG.Fp1.15 | 1 | 1 | 1 |

Per unit change in regressor

Per change in regressor over entire range is as follows.

**Table 4.** Range Odds Ratios

| Term | Odds Ratio | Lower 95% | Upper 95% |
| --- | --- | --- | --- |
| EEG.Fp1 | 1 | 1 | 1 |
| EEG.Fp1.1 | 1 | 1 | 1 |
| EEG.Fp1.2 | 1 | 1 | 1 |
| EEG.Fp1.3 | 1 | 1 | 1 |
| EEG.Fp1.4 | 1 | 1 | 1 |
| EEG.Fp1.5 | 18.564673 | 1.0449909 | 329.80868 |
| EEG.Fp1.7 | 1 | 1 | 1 |
| EEG.Fp1.8 | 1 | 1 | 1 |
| EEG.Fp1.9 | 1 | 1 | 1 |
| EEG.Fp1.10 | 0.0316477 | 0.0015223 | 0.6579313 |
| EEG.Fp1.11 | 1 | 1 | 1 |
| EEG.Fp1.12 | 1 | 1 | 1 |
| EEG.Fp1.13 | 1 | 1 | 1 |
| EEG.Fp1.14 | 0.0211304 | 0.00068 | 0.6566062 |
| EEG.Fp1.15 | 1 | 1 | 1 |

Per change in regressor over entire range

### 4. Classification models

The features extracted by LASSO are fed into machine learning kernels in MATLAB with 7 fold cross validation and the following results were obtained:

#### 1. Fine Gaussian SVM

Fine gaussian SVM model showing confusion matrix and an AUROC of **73%**.

#### 2. Decision Tree

Decision tree model showing confusion matrix and an AUROC of **80%**.

### Summary of kernel models

Decision tree had a better accuracy than Fine Gaussian SVM as shown in the table.

**Table 5.** Summary of kernel model.

| Algorithm/model | Accuracy | Precision | Recall | F1-measure |
| --- | --- | --- | --- | --- |
| 1. Fine Gaussian SVM | 0.73 | 0.79 | 0.60 | 0.68 |
| 2. Decision Tree | 0.78 | 0.77 | 0.80 | 0.78 |

## Discussion

This work has elucidated the chaotic nature of the brain waves in the face of serial cognitive workload and discrimination of the ‘good’ and ‘bad’ performers when faced with increasing cognitive load. The broader objective of this work is to develop biomarkers which can discriminate good performers from bad performers involved in a n-back task. We found that theta power spectral density, Higuchi’s fractal dimension and standard deviation of the signals are the biomarkers having a good discriminatory power by machine learning.

Theta band oscillation directly reflected cognitive processes. This finding was much congruent with the papers which reported that alpha and theta oscillations were reflective of cognitive and memory performance^23–25^. It was also found to be in congruence with the findings described by Molnar et al^26^. Specifically, good group had lower entropy of theta and beta band in frontal lobe when compared to the bad group. This implies that the complexity of the brain dynamics is less in the frontal, and more in the occipital and temporal groups in complex cognitive processes. In other words, higher the mental workload on the memory, which has happened with the good group, lower is the entropy of the EEG signals in the frontal and higher in the occipital and temporal lobes. Thus, a decreased value of entropy with increased task load implies higher predictability and less irregularity in the brain activity. This finding is supported by Zarjam et al who showed that entropy decreases with increase in cognitive load mainly in the frontal lobe, but it was more prominent in the delta sub band and they had measured approximate entropy and spectral entropy^20^. Both these studies showed that brain behaves in a more focused and regular manner when performing more difficult tasks. Further, Entropy is also found to be correlated with fractal dimensions positively. In other words, Fractal dimension behaved similarly to that of entropy. Good group had lower entropy, and FD of theta and beta band in frontal lobe than bad group, while good group had higher entropy and FD of theta and beta in occipital lobe and temporal lobe. While Wang et al reported a stark contrast study that Fractal dimension increased with increased cognitive load mainly in the frontal lobe^27^, beyond our understanding. PSDs of theta and beta bands were higher in frontal but lower in occipital lobe when compared to bad group. This shows that higher PSD correlates with higher mental workload in frontal lobe. Zafar et al showed the involvement of delta oscillations in occipital region during cognitive tasks and they also showed that during the cognitive task the delta band has a different behaviour compared to the rest task^28^. But they didn’t classify the task group into good and bad performers. This could be due to the fact that their experiment was done with the subjects’ eyes closed.

Due to the processed non-linear nature and a good generalizability of the feature set, the polynomial kernel function was used in our fine gaussian SVM model, as such approaches have been reported in similar situations. The feature had a better predictive capacity with a good area under ROC to discriminate rest states from task states. Decision tree produced the highest accuracy **78%**. Attallah et al used the same dataset to classify mental stress states and non-stress states using Principal Component analysis (PCA)^29^. PCA does feature reduction from such electrodes to lower model complexity, where the optimal number of principal components is examined using sequential forward procedure. It also examines the minimum number of electrodes placed on the site which has greater impact on stress detection. They showed that using only 58 and 15 principal components, the accuracy of detecting stress reached **100%** and **99**.**8%** using cubic SVM and KNN classifiers, respectively, whereas our study showed an accuracy of **73%** with SVM. But PCA’s drawback lies on the fact that it causes overfitting in high dimensional datasets as EEG datasets as mentioned earlier when compared to the LASSO model which we have used. F. C. Galan and C. R. Beal^54^ did a piecemeal analysis of the entire signal breaking it off into first or the last 20 seconds but the prediction accuracy to predict a correct or incorrect outcome was **77%** and **79%** by SVM respectively which didn’t yield a significant difference in accuracy as did ours. The SVM accuracy for easy vs hard problems for both predictions using engagement and workload yielded an accuracy of **73%** which was significantly lower than ours, **79%**. This might be hypothesized due to the fact that we had studied the continuous time series changes of the signals rather than a cross section using non-linear features as an add on to the linear features for classification and estimation of the workload. Winnie K. Y. So et al^55^ have identified the key EEG spectral feature associated with mental workload and explored the feasibility to classify different levels of mental workload from EEG spectral features using SVM. The accuracy of their mental workload classification could reach **65%–75%** which was lesser than our classification accuracy. This might be due to the fact that we have included non-linear features in our analysis too or that the sequential feature selection methods adopted could have boosted the classification accuracy performance.

A limitation of the study is that for each arithmetic calculation load, the amount of change in the feature could not be quantified because of the gross variation for each subject in the calculation they could perform. We also didn’t perform any analysis of the transition state from the rest to task state as it is very abrupt and the transition could be easily delineated. In future, this study can be potentially extrapolated to subjects with dementia or minimal cognitive impairment and will be able to detect such degenerative processes at earlier stages and can help to understand the progression of these diseases as far as electrophysiology is concerned. This study also ensures productivity in task completion and applications in ergonomics.

### Conclusion

Frontal lobe is affected more widely and deeply during mental arithmetic task cognitive load. The hybrid machine learning model, SMORASO-DT (SMOte + Random forest + lASso-Decision Tree), differentiates good and bad performers during task state with an accuracy of 79%.

## Methods

The study is an analysis of the data originally collected from physionet.org published by Zyma et al. Briefly the methodology of the study is described herein.

### Participants

The dataset was obtained from the PhysioNet “EEG During Mental Arithmetic Tasks” database contributed by contributed by Igor Zyma, Sergii Tukaev, and Ivan Seleznov, National Technical University of Ukraine “Igor Sikorsky Kyiv Polytechnic Institute”, Department of Electronic Engineering^30,31^.(Zyma et al, Goldberger et al physiobank). The original article from which the data has been obtained has explained this in detail.

Totally, 66 healthy right-handed volunteers (47 women and 19 men) were initially involved in the study. All participants are 1st–3rd year students of the Taras Shevchenko National University of Kyiv (Educational and Scientific Centre “Institute of Biology and Medicine” and Faculty of Psychology) aged 18 to 26 years (Mean = 18.6 years, Standard Deviation (SD) = 0.87 years). The details of the participants are described in the original article on which the analysis is performed on.

The participants were included if they had normal or corrected-to-normal visual acuity, normal color vision, and had no clinical manifestations of mental or cognitive impairment or verbal or non-verbal learning disabilities. Exclusion criteria were the use of psychoactive medication, psychiatric or neurological complaints and drug or alcohol addiction.

The study was approved by the Bioethics Commission of Educational and Scientific Centre “Institute of Biology and Medicine”, Taras Shevchenko National University of Kyiv (Conclusion from 15 August 2018, project title “Detrended fluctuation analysis of activation re-arrangement in EEG dynamics during cognitive workload”). Each subject signed written informed consent following the World Medical Association (WMA) declaration of Helsinki of 1975 (http://www.wma.net/Data 2019, 4, 14 5 of 6 en/30publications/10policies/b3/), revised in 2008, the Declaration of Principles on Tolerance (28th session of the General Conference of UNESCO, Paris, 16 November 1995), the Convention for the protection of Human Rights and Dignity of the Human Being with regard to the Application of Biology and Medicine: Convention on Human Rights and Biomedicine (Oviedo, 4 April 1997). If the subject is below 18 years, then informed consent was obtained from parents/legally authorized representatives.

## Apparatus/materials

The entire analysis is summated in Figure 8. Neurocom monopolar EEG 23-channel system (Ukraine, XAI-MEDICA) was used to obtain the recording. Silver/silver chloride electrodes were placed on the scalp at symmetrical anterior frontal (Fp1, Fp2), frontal (F3, F4, Fz, F7, F8), central (C3, C4, Cz) parietal (P3, P4, Pz), occipital (O1, O2), and temporal (T3, T4, T5, T6) recording sites as per the ***10-20 International*** System of Electrode Placement. All electrodes were referenced to the interconnected ear reference electrodes. The inter-electrode impedance was below 5 kΩ and sample rate 500 Hz per channel.

**Figure 8.**
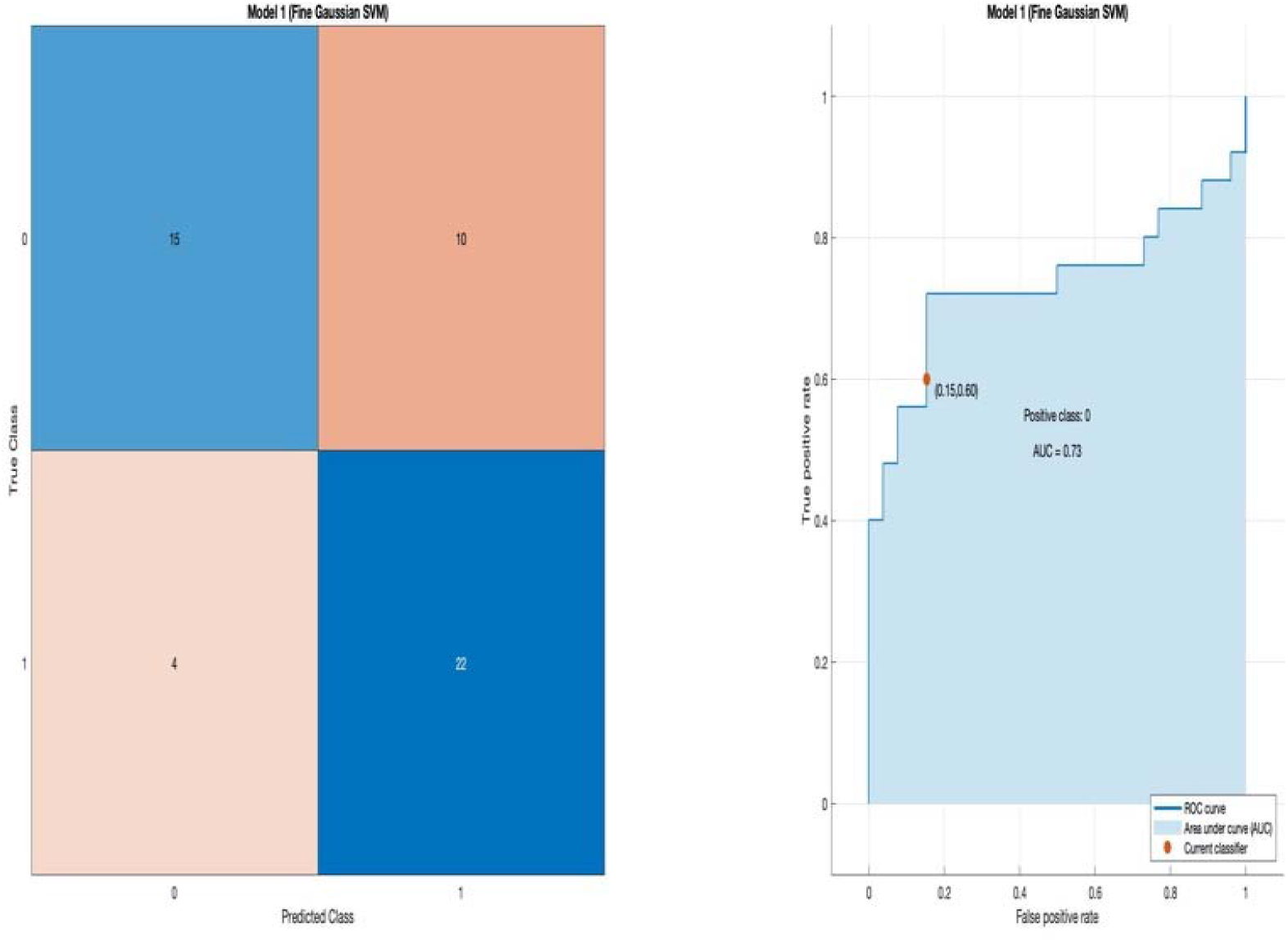
Fine Gaussian SVM showing the confusion matrix and an AUROC of **73%**.

Each recording consists of an artefact-free EEG segments of 180 s for resting state and 60 s for task state. 30 of the 66 initial participants were excluded from the database due to poor EEG quality by a board certified electro neurophysiologist, hence the final number 36 subjects.

### Procedure

Tasks in this study involved the serial subtraction of two numbers. Each trial started with the oral communication of the 4-digit (minuend) and 2-digit (subtrahend) numbers (e.g., 7456 and 45, 2343 and 48, etc.). Serial subtraction during 15 min is considered to be a psychosocial stress^32^. In this way, our study design required intensive cognitive activity from the subjects. Intensive mental load is accompanied by a change in the emotional background when the subject makes additional effort to resolve tasks, so that they can talk about evoked emotions in this case. During EEG recording, the participants sat in a dark soundproof chamber, comfortably reclined in an armchair. Serial subtraction is a composite measure of auditory attention/ concentration, mental tracking, and computation. The experiment is divided into three phases: the adaptation phase for 3 min, the resting phase for 3 min and finally the task phase/ the mental counting phase for 4 min. To avoid all the distractors, before the experiment is begun, participants were instructed to relax during the rest state. After 3 minutes of adaptation to experimental conditions, EEG registration of the rest state with closed eyes was made (over the next 3 minutes). Then the participants performed a mental arithmetic task—serial subtraction—for 4 min. For each participant, the last number reached from the initial 4-digit number is subtracted and a mental arithmetic score is obtained. If the score was an exact multiple of a corresponding 2-digit subtrahend, then the task performance was considered accurate. Then the mentally calculated subtraction was compared across subjects. The participant had successfully engaged in the task if their reported result did not differ by more than 20% from the correct value and was labelled as good.

Participants were interviewed about their strategies and experience after the experiments. These periods were selected since the task performance strategy is being formed simultaneously as the task is executed, and the emotional state of the participants is changing considerably due to intellectual overload.

### Data processing and analysis

EEG Pre-processing. EEG data were first exported to EEGLAB^33^ then analysed mainly using MATLAB (R2018b). The length of rest state EEG signal is 180 seconds and the length of mental arithmetic EEG signal is 60 seconds. They were divided into 10-seconds epochs. However, the rest state EEG signal of subject 04 and subject 31 are shorter than other. Therefore, there are 851 epochs, including 635 rest epochs and 216 task epochs.

A high-pass and a low pass filter with cut-off frequency 0.5 Hz and 45 Hz, respectively, 50 Hz power line notch filter and 0.3 s time constant of the amplification tract were used. The Independent Component Analysis (ICA) method was used to eliminate the artifacts (eyes, muscle and cardiac overlapping of the cardiac pulsation). The wave and power spectral density (PSD) of clean pre-processed data is shown in Figure 9. There are 19 channels, including FP1, FP2, F3, F4, F7, F8, T3, T4, C3, C4, T5, T6, P3, P4, O1, O2, Fz, Cz and Pz.

**Figure 9.**
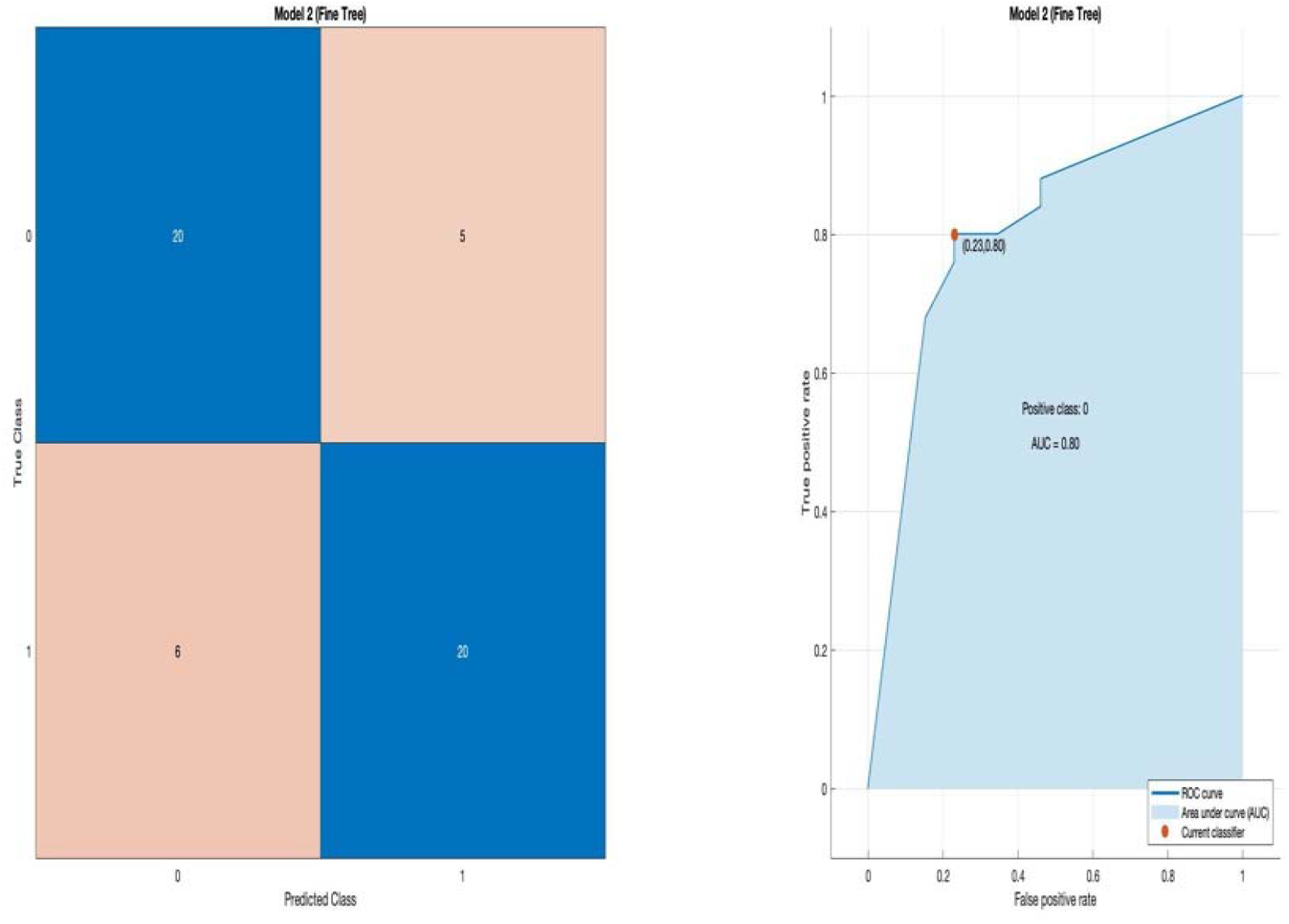
Decision tree model showing confusion matrix and an AUROC of 80%.

**Figure 10.**
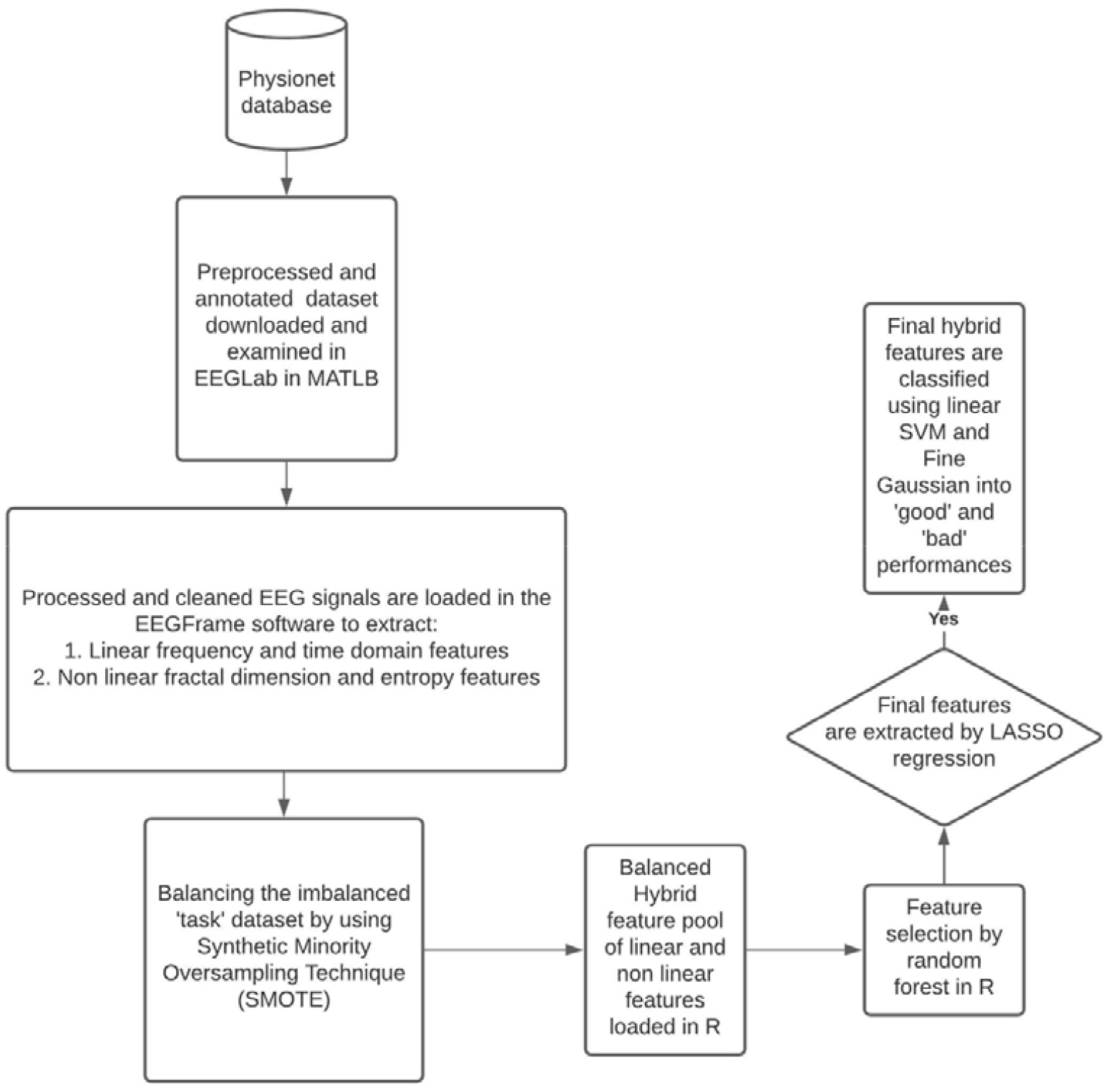
Workflow of the analysis.

### Decomposition

The EEG data was decomposed into several frequency bands (delta, theta, alpha, beta and low gamma) based on discrete wavelet 5 level wavelet db2 carried out in MATLAB (R2018b) (MathWorks Inc., Natick, MA).

According to the following formula^34^ (Daubechies), the decomposition was carried out and the results were obtained as shown in Table 6. and Figure 11.

**Table 6.** Decomposition of the EEG data: Frequency range of corresponding bands.

| Frequency Range | Name |
| --- | --- |
| 1-4Hz | delta |
| 4-8Hz | theta |
| 8-14Hz | alpha |
| 14-30Hz | beta |
| 30-100Hz | Low gamma |

**Figure 11.**
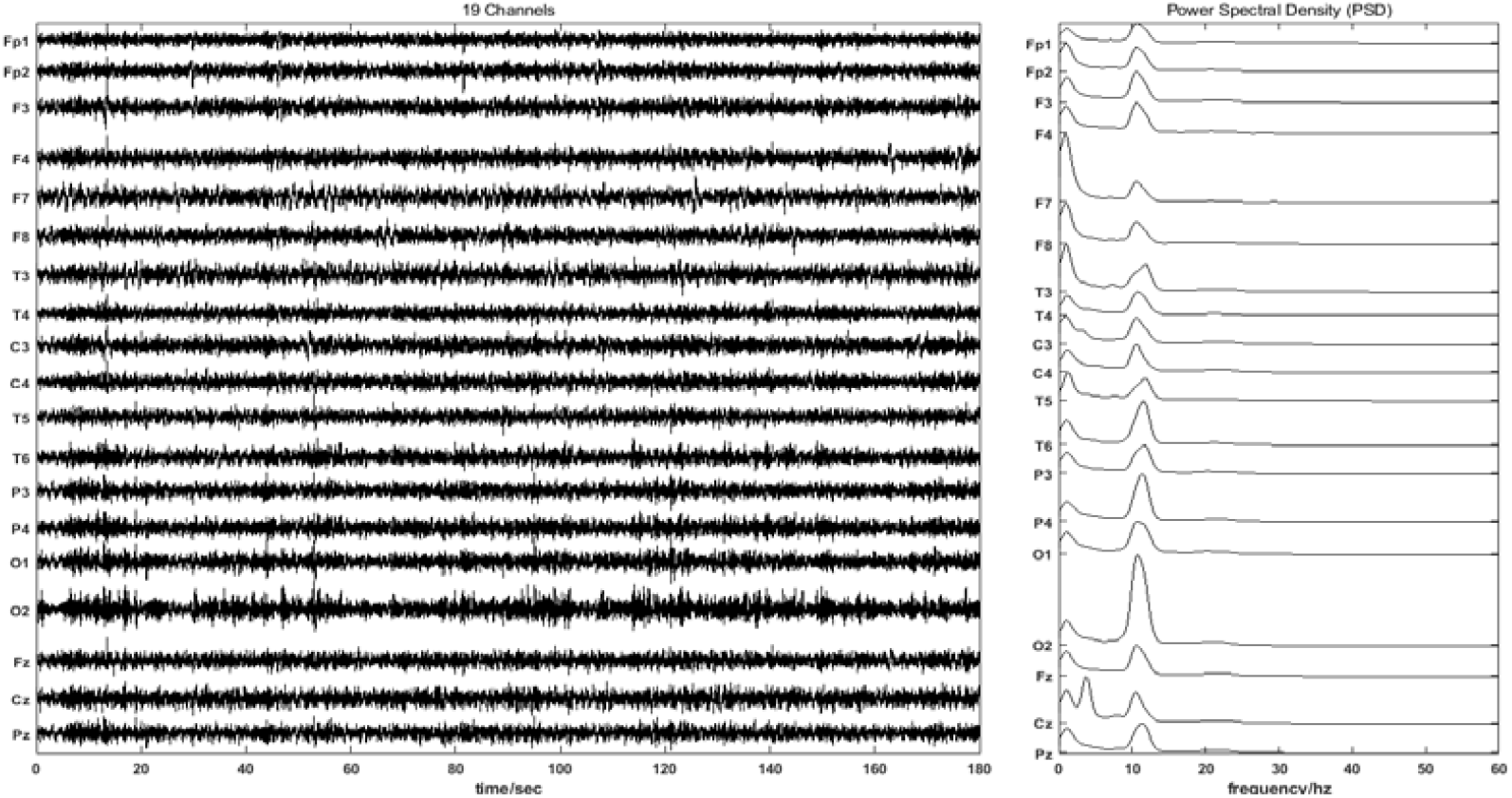
Processed signals of 19 channels with the estimated corresponding power spectral densities.

**Figure 12.**
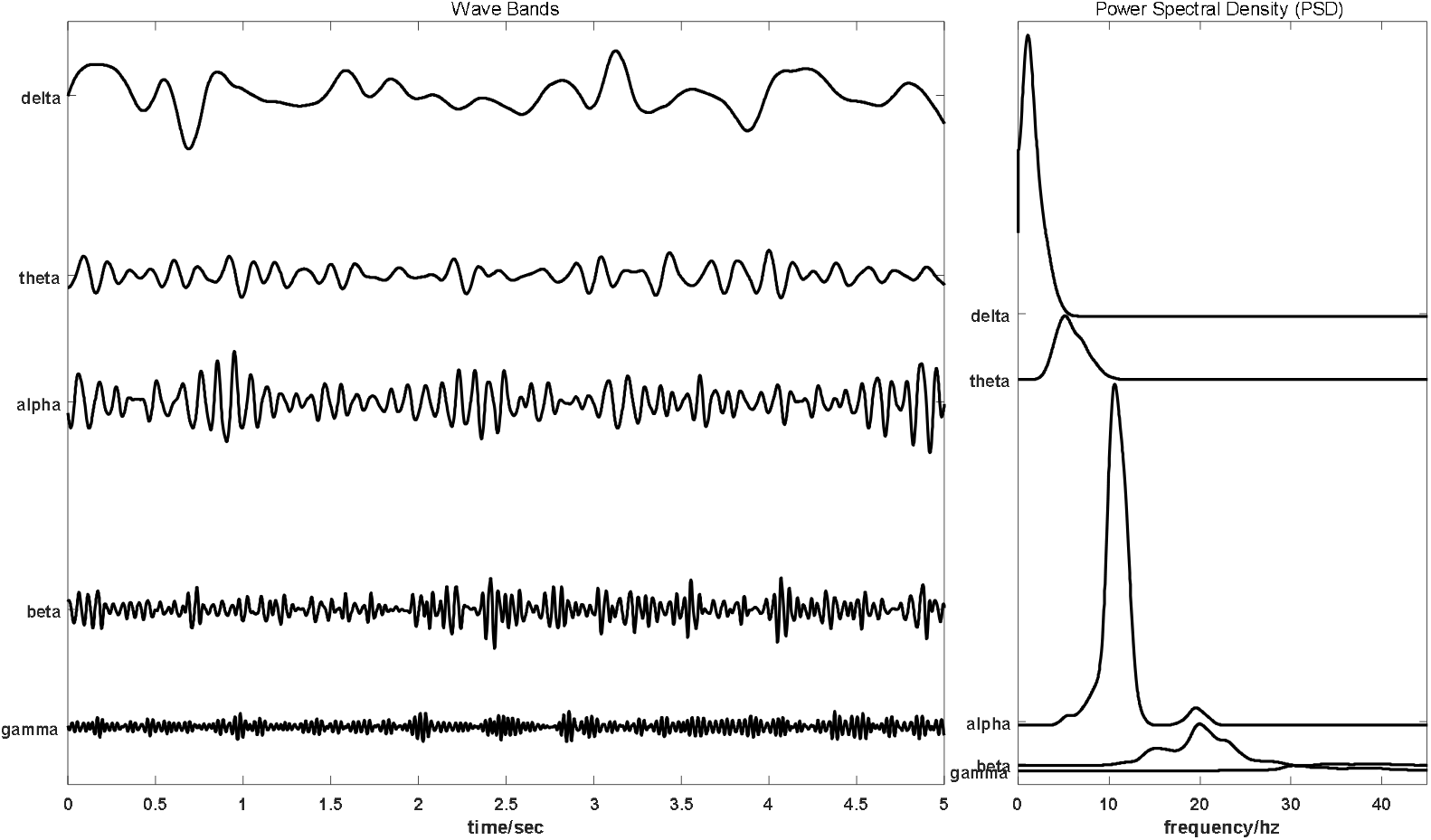
Decomposition of the EEG data into different frequency bands.

### Feature Extraction

In this study, the following features were used on each EEG channel of the subjects during task phase using EEGFrame software(http://www.zemris.fer.hr/~ajovic/eegframe/eegframe.html).

**Table 7.**
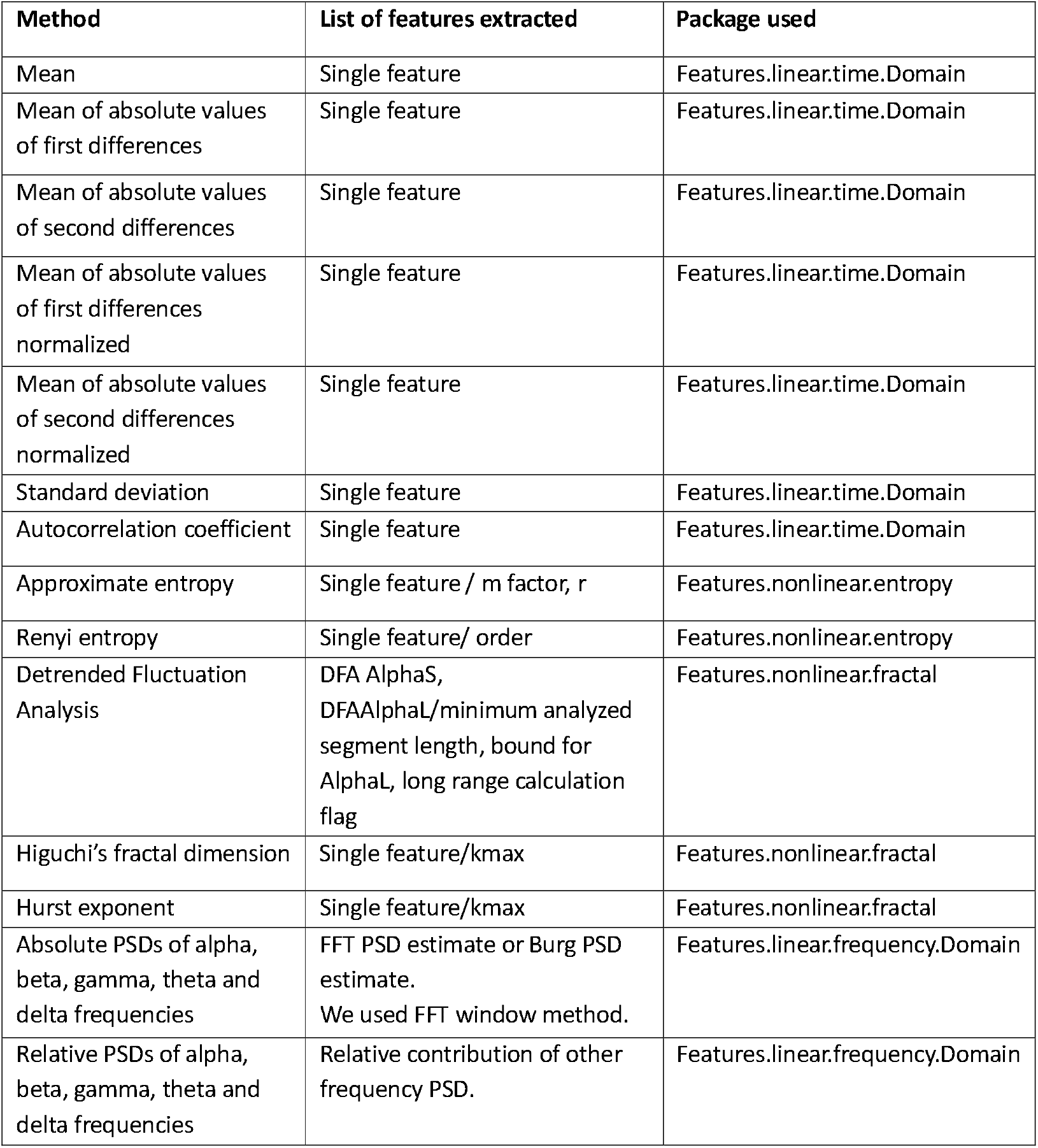
List of features extracted, packages used and methods adopted by EEGFrame software.

### Power spectral density

*Welch’s method*^35^ was used for estimating power spectra. It is carried out by dividing the time signal into successive blocks, forming the periodogram for each block, and averaging.

The **m**th windowed, zero-padded frame from the signal **x** is denoted by

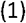

where *R* is defined as the window *hop size*, and let *K* denote the number of available frames. Then the periodogram of the **m** th block is given by

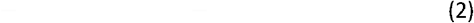

as before, and the Welch estimate of the power spectral density is given by

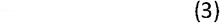

In other words, it’s just an average of periodograms across time. When **w(n)** is the rectangular window, the periodograms are formed from non-overlapping successive blocks of data.

#### Mean

Arithmetic Mean is the mathematical representation of the typical value of a set of data, computed as the sum of all the numbers in the dataset divided by the size of the dataset. If the sample space is {x_1_, x_2_, x_3_….x_n_}, then the arithmetic mean µ_x_ is defined as the means of the raw signals

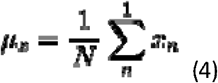

#### Standard Deviation

Standard deviation measures the degree of variation or ‘dispersion’ which exists from the mean. A low standard deviation indicates that the data points tend to be very close to the mean, whereas the high standard deviation indicates that the data points are spread out over a large range of values. If X is a random variable with mean value µ, then the standard deviation of X is:

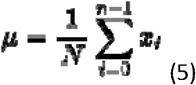

#### Sample entropy

Sample entropy is a modification of approximate entropy.^36^ We have an imfN=xi=(x1,x2,x3,…,xN) i=1,2,3,…,N and use a time interval to reconstruct series Xm(i)=(xi,xi+1,xi+2,…,xi+m−1) i=1,2,3,…,N−m+1. The length of sequence is *m*. The distance function of two sequences is d[Xm(i),Xm(j)]. If the embedding dimension is *m*, tolerance is *r* and number of data points is *N*, SE is expressed as:

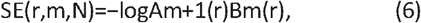

where, Bm(r) is the number of template vector pairs having d[Xm(i),Xm(j)]<r and represents the similarity between two sequences of length *m*, Am+1(r) is the number of template vector pairs having d[Xm+1(i),Xm+1(j)]<r and represents the similarity between two sequences of length *m* + 1.

#### Fractal dimension

Fractal dimension as one of the chaos measurement tools is widely utilized in the determination of chaotic behaviour of signals. We have used the box-counting method for the FD calculation. In this method, a network of squares is created on the contour, and the number of squares in this network that includes a part of the curve is calculated. The size of the squares changes, then, the occupied squares are counted again. A point is obtained by calculating the logarithm of the number of counted squares and the logarithm for the reduction coefficient. New points are obtained by reducing the size of the network, which is connected in a curve. Calculating the slope of this curve indicates the FD. Ahmadlou et al compared two main algorithms for calculating fractal dimension from EEG. HFD was shown to provide better discrimination (91.3%) compared to Katz’s Fractal Dimension^37^. In this paper we have used the Higuchi’s FD due to its high level of accuracy^38^.

Equation (9) describes how the self-similarity dimension (D), the number of self-similar pieces (a) and the reduction factor (1/S) are related together.

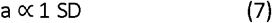

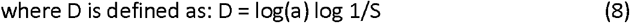

D can be calculated by estimating the slope of the approximated line for the plot of log (a) vs. log (1/s).

#### Hurst exponent

If the time series x(t) repeats the same statistical features across multiple temporal scales it is said to be self-similar.^39^ Hence, scaling along the time axis by a factor of “a” requires a rescaling along the signal amplitude axis by a factor of a^H^ ; which is, x(at) = a^H^ x (t) for all t > 0, a > 0 and H > 0. The Hurst exponent is a measure of quantifying self-similarity in signals.

Rescaled range analysis is used to estimate the Hurst exponent of an N-long time series x.^40^

The average rescaled range is modelled as a power law function over *n* whose asymptotic behaviour represents the Hurst exponent, H.

*H* is estimated as the slope of the logarithmic plot of the rescaled ranges versus *ln*(*n*). Rescaled range analysis quantifies the fluctuations of a signal around its stable mean.

### Approximate entropy

ApEn involves the following parameters: the vector length m, the “filter factor” r, and the number of data points N. ApEn measures the logarithmic likelihood that sets of patterns that are close for m-observations remain close on the next incremental comparisons.^41^ ApEn characterizes how different segments of the signal with similar recent histories remain similar in the future. Insofar as ApEn decreases, the complexity of the signal is low and determinism is high. As described by Pincus,^41^ the ApEn is computed as:

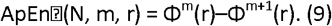

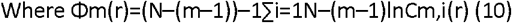

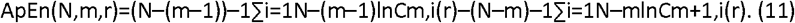

### Detrended fluctuation analysis

DFA has been used commonly to quantify the long-range correlation embedded in a non-stationary time series^42,43^. The standard procedure of the DFA:

1. Observed time series {*x*_i_} after subtracting the mean from each data point (a) is integrated.
2. The integrated time series is divided into equal-sized, non-overlapping segments of length *n* samples.
3. In each segment, the mean-square-deviation from the least-squares polynomial fit of degree *k* is calculated. Depending on the polynomial degree *k*, the method is referred to as *k*th-order DFA or DFA*k*, in which non-stationary trends approximated by polynomial functions of degree (*k*-1) are removed from {*x*_i_}. The mean-square-deviations are then averaged over all segments and its square root *F*(*n*) is calculated. This computation steps (2) and (3) are repeated over multiple time scales (window sizes) to characterize the relationship between *F*(*n*) and *n*. The power-law scaling range is indicated by a linear relationship on a log-log plot of *F*(*n*) as a function of *n*.

### SMOTE

Once the features are extracted from the subjects, the imbalanced features are balanced by using the SMOTE technique in MATLAB. Bad group has 12 subjects but the good group has 24 subjects. This makes the data set grossly imbalanced. This function synthesizes new observations based on existing (input) data, and a k-nearest neighbour approach. If multiple classes are given as input, only neighbours within the same class are considered. The function^44^being

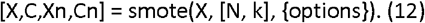

Wherein,

Input:

% X: Original dataset. Each row is an observation, and each column a different feature.

% N: Amount of oversampling (Default: 1 = 100% doubles the observations)

If ‘Class’ is set, then N can be a vector with number of elements equal to the number of classes. Elements of N should be sorted according to the sorted unique classes. If N is empty, then ‘balancing’ will occur, synthesizing minority classes to the same number as the majority class.

% k: Number of nearest neighbours to consider. If ‘Class’ is set, then k can be a vector with number of elements equal to the number of classes. Elements of k should be sorted according to the sorted unique classes.

Output:

% X: Complete dataset (original and synthesized)

% C: Classes of complete dataset

% Xn: Synthesized observations

% Cn: Classes of synthesized observations

We have developed a hybrid machine learning model which can differentiate good and bad performers during task state which is the first of its kind. We used a two-step approach: 1. robust variable selection using the random forest approach followed by 2. adaptive lasso methods that incorporate both dimensionality reduction and quantification of the degree of association between the features and the outcome of interest. We used the RF approach as it effectively handles datasets with many more variables than subjects, captures nonlinear relationships between predictor variables, effectively handles missing values, and produces more robust results that are insensitive to outlier effects. The importance scores of features can be misleading when the features are similar to each other. When there are more redundant features, the importance of each feature becomes even smaller. This might not affect the performance accuracy but could be misleading in interpretation. The solution would be to regularize the trees of the random forest. In the tree building process, regularization memorizes the features used in previous tree nodes, and prefer these features in splitting future tree nodes, therefore avoiding redundant features in the trees.

### Feature selection by Random forest

#### Feature Selection

Random forest [RF] learning algorithm due to its non-parametric assumptions and bootstrapping methods, used to identify which of the features were most useful for discriminating between the good and bad performers of the arithmetic task. The mean decrease in Gini Index (MDG) and accuracy decrease of the RF upon removal of the variable, were used in identifying the top ten features useful in the discrimination. For the development of an assessment tool that maximizes the potential of these features in differentiating good and bad performers, multivariable logistic regression models were created using these top ten features.

### Filtering out the features further with LASSO regression

Pattern recognition is an important step in accurately classifying EEG signals in Brain Computer Interface(BCI). Many EEG classification algorithms have been advocated, including logistic regression, support vector machine^45^, decision tree^46,47^, and convolutional neural networks^48,49^. These methods mostly aim to find a direct classification model without involving any sparse processing. However, as EEG data is a high dimensional data with a large feature set and a disproportionately small sample size, these methods are prone to over-fitting or low precision. The main approach to overcome this limitation is regularization, currently represented by the *L*_0_, Lasso (*L*_1_), Ridge Regression, and Elastic Net methods^50–53^. *L*_0_ penalty is used in the case of sparseness, but this method involves an NP-hard problem. Hence, the Lasso (*L*_1_) penalty is most often used. Adaptive lasso is a modified version of lasso that was developed to address the limitations of lasso. Using a flexible weighting scheme, the adaptive lasso applies different amounts of shrinkages to different coefficients, whereas lasso applies the same penalty to every regression coefficient which may induce potential bias. It combines the good features of both subset selection and ridge regression. Adaptive lasso penalizes the weighted L_1_-norm of the regression coefficients, and the coefficient estimates are defined as:

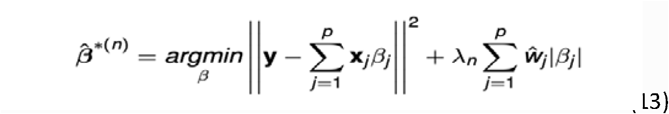

where ***w*** is a weighing vector with =||| ^ |||− wj=|β^jinitial|−γ.

The LASSO regression shrinkage parameter was determined using 10-fold cross-validation within each data set. Receiver Operating Characteristic (ROC) curves were used to estimate the extent to which these features were useful in the classification. Features which were statistically significant which were extracted are : Standard deviation, Higuchi Fractal Dimension and Theta power spectral density.

### Classification models

#### Classified Forecast

In the task of classification and prediction, we compare the effect of the model used in this paper with the following benchmark models. (1) *Decision Tree* (*DT*) *Algorithm*. It is a method of approaching the value of discrete function, which is a typical classification method. Firstly, the data is processed, the readable rules and decision trees are generated by using an induction algorithm, and then the new data is analysed by using decision. In essence, the decision tree is a process of data classification through a series of rules. (2) *Support Vector Machines* (SVM) *Algorithm*. SVM finds a hyperplane to divide the data into one class and other classes, which is a two-class classification model. The separation interval is the largest and different from the perceptron. All models are executed and obtained using MATLAB.

### Prediction Scheme

In order to evaluate the prediction efficiency of the classification prediction task, we use common evaluation indicators: accuracy, precision, recall, and F1-measure. Specifically, the definitions of accuracy, precision, recall, and F1-measure are as follows:

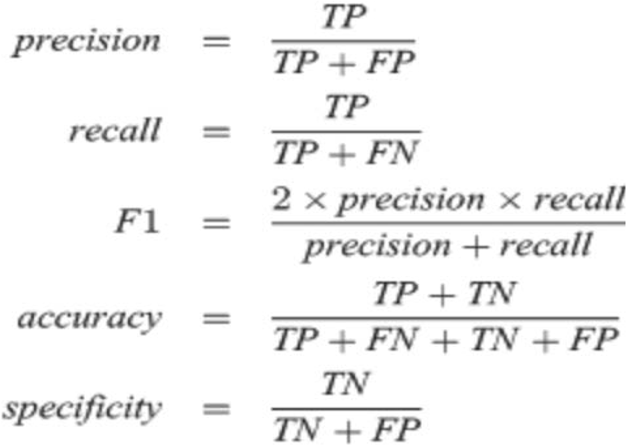

True positive (TP) is the number of correctly performed tasks which are good, False positive (FP) is the number incorrectly recognized as good performers, True Negative (TN) is the number of correctly recognized good performers, and False Negative (FN) is the number of incorrectly recognized open eyes. The accuracy rate is the ratio of the number of samples correctly classified by the classifier to the total number of samples in a given test data set, the accuracy is the ratio of the number of correctly predicted samples in all predictions, the recall rate is the correctly predicted positive samples in all actual prediction proportion, and the F1-measure is the harmonic average of the exact value and the recall rate. These four evaluation schemes help us compare the efficiency of different classification models in the EEG data set for mental arithmetic task performance, and the bigger the results of the two evaluation methods, the closer the prediction results to the actual situation.

In this paper, the accuracy, precision, recall rate, and F1-measure are used to evaluate the prediction efficiency of different models.

### Statistical software

All analyses were carried out in MATLAB (R2018b) (MathWorks Inc., Natick, MA)^54^, EEGFrame^55^ and R studio^56^. All reported statistical tests in the present study are two-sided tests wherever applicable. The significant difference was defined as the p-value < 0.05. Results were reported following the ‘Transparent Reporting of a Multivariable Prediction Model for Individual Prognosis or Diagnosis’ (TRIPOD)^57^ and ‘Guidelines for Developing and Reporting Machine Learning Predictive Models in Biomedical Research’^58^ statements.

## Data Availability

The data analysed in the study are available to the reader on reasonable request.

https://physionet.org/content/eegmat/1.0.0/

## Author contributions

S.K.R conceived the experiment. S.K.R. and B.D. analyzed the data. S.G. and A.R. analyzed the results. S.G. wrote the manuscript. All authors reviewed the manuscript.

